# A turning point in COVID-19 severity and fatality during the pandemic: A national cohort study in Qatar

**DOI:** 10.1101/2023.05.28.23290641

**Authors:** Hiam Chemaitelly, Houssein H. Ayoub, Jeremy Samuel Faust, Peter Coyle, Patrick Tang, Mohammad R. Hasan, Hadi M. Yassine, Hebah A. Al-Khatib, Asmaa A. Al Thani, Zaina Al-Kanaani, Einas Al-Kuwari, Andrew Jeremijenko, Anvar Hassan Kaleeckal, Ali Nizar Latif, Riyazuddin Mohammad Shaik, Hanan F. Abdul-Rahim, Gheyath K. Nasrallah, Mohamed Ghaith Al-Kuwari, Adeel A. Butt, Hamad Eid Al-Romaihi, Mohamed H. Al-Thani, Abdullatif Al-Khal, Roberto Bertollini, Laith J. Abu-Raddad

**Affiliations:** Infectious Disease Epidemiology Group, Weill Cornell Medicine-Qatar, Cornell University, Doha, Qatar; World Health Organization Collaborating Centre for Disease Epidemiology Analytics on HIV/AIDS, Sexually Transmitted Infections, and Viral Hepatitis, Weill Cornell Medicine–Qatar, Cornell University, Qatar Foundation – Education City, Doha, Qatar; Department of Population Health Sciences, Weill Cornell Medicine, Cornell University, New York, New York, USA; Mathematics Program, Department of Mathematics, Statistics, and Physics, College of Arts and Sciences, Qatar University, Doha, Qatar; Department of Emergency Medicine, Brigham and Women’s Hospital, Boston, Massachusetts, USA; Hamad Medical Corporation, Doha, Qatar; Biomedical Research Center, QU Health, Qatar University, Doha, Qatar; Wellcome-Wolfson Institute for Experimental Medicine, Queens University, Belfast, United Kingdom; Department of Pathology, Sidra Medicine, Doha, Qatar; Department of Biomedical Science, College of Health Sciences, QU Health, Qatar University, Doha, Qatar; Department of Public Health, College of Health Sciences, QU Health, Qatar University, Doha, Qatar; Primary Health Care Corporation, Doha, Qatar; Department of Medicine, Weill Cornell Medicine, Cornell University, New York, New York, USA; Ministry of Public Health, Doha, Qatar; College of Health and Life Sciences, Hamad bin Khalifa University, Doha, Qatar

**Keywords:** COVID-19, severe, critical, fatal, acute-care, ICU, cohort study, Qatar, epidemiology

## Abstract

**Background:** This study assessed the evolution of COVID-19 severity and fatality by utilizing rigorous and standardized criteria that were consistently applied throughout the pandemic in Qatar.

**Methods:** A national cohort study was conducted on Qataris, using data on COVID-19 acute-care and ICU hospitalizations, as well as severe, critical, and fatal COVID-19 cases classified according to the World Health Organization criteria.

**Results:** The cumulative incidence of severe, critical, or fatal COVID-19 after 3.14 years of follow-up was 0.45% (95% CI: 0.43-0.47%). The incidence rate for severe, critical, or fatal COVID-19 throughout the pandemic was 1.43 (95% CI: 1.35-1.50) per 1,000 person-years. In the pre-omicron phase, first omicron wave, and combined phases, it was 2.01 (95% CI: 1.90-2.13), 3.70 (95% CI: 3.25-4.22), and 2.18 (95% CI: 2.07-2.30) per 1,000 person-years, respectively. The post-first omicron phase saw a drastic drop to 0.10 (95% CI: 0.08-0.14) per 1,000 person-years, a 95.4% reduction. Among all severe, critical, and fatal cases, 99.5% occurred during the primary infection. The cumulative incidence of fatal COVID-19 was 0.042% (95% CI: 0.036-0.050%), with an incidence rate of 0.13 (95% CI: 0.11-0.16) per 1,000 person-years. In the post-first omicron phase, the incidence rate of fatal COVID-19 decreased by 90.0% compared to earlier stages. Both severity and fatality exhibited an exponential increase with age and a linear increase with the number of coexisting conditions.

**Conclusions:** The conclusion of the first omicron wave was a turning point in the severity of the pandemic. While vaccination and enhanced case management reduced severity gradually, the rapid accumulation of natural immunity during the initial omicron wave appears to have played the crucial role in driving this shift in severity.

## Introduction

The coronavirus disease 2019 (COVID-19) pandemic, caused by the severe acute respiratory syndrome coronavirus 2 (SARS-CoV-2), has resulted in significant morbidity and mortality globally.^1–4^ The pandemic has also caused extensive economic losses and societal disruptions due to the necessary implementation of social and physical distancing measures aimed at reducing virus transmission.^5^ The need for future public health interventions and restrictions will depend on the ongoing evolution of the virus, its disease severity, and the effectiveness of current and future interventions.

The World Health Organization (WHO) has established a specific classification system for COVID-19 case severity,^6^ criticality,^6^ and fatality.^7^ Qatar appears to be the only country to have consistently implemented this standardized WHO classification at a national level to assess the severity of COVID-19 cases from the pandemic onset to the present. Trained medical personnel evaluate the severity of COVID-19 cases using a national protocol that applies to every hospitalized COVID-19 patient.^1, 8^ Importantly, COVID-19-associated hospitalizations are not used as a proxy for COVID-19 severity, as these have limitations in accurately capturing the true severity of COVID-19.^9, 10^ This presents a special opportunity to investigate the evolution of COVID-19 severity from the start of the pandemic to the present time, using rigorous and standardized criteria in a national population cohort.

The objective of this study was to examine how COVID-19 severity changed over the course of the pandemic in response to subsequent waves of infection, increasing population immunity,^11^ and the emergence of new viral variants. The study analyzed the incidence of COVID-19 acute-care hospitalizations, COVID-19 intensive care unit (ICU) hospitalizations, as well as the incidence of severe,^6^ critical,^6^ and fatal^7^ COVID-19 cases, according to the WHO classification, in the national cohort of Qatari citizens.

Qatar’s population includes Qataris and a large expatriate workforce.^12^ A considerable proportion of the expatriate population consists of healthy men who are craft and manual workers aged 20-49,^13, 14^ a population possibly affected by the healthy worker effect.^15, 16^ As a result, they may not represent a natural national population.^16^ In contrast, Qatari citizens represent a typical national population, including both healthy and unhealthy individuals, making them relevant for assessing the severity of SARS-CoV-2 infections in this study.

## Methods

### Study population and data sources

This study assessed the severity of COVID-19 in the national cohort of Qatari citizens from February 28, 2020, which marks the earliest record of a SARS-CoV-2-positive test in Qatar, to April 21, 2023. The study utilized the national, federated databases for COVID-19 laboratory testing, vaccination, hospitalization, and death, obtained from the integrated, nationwide, digital-health information platform (Section S1 of the Supplementary Appendix). The databases contain SARS-CoV-2-related data with no missing information since the pandemic’s onset, including all polymerase chain reaction (PCR) tests and, from January 5, 2022, all medically supervised rapid antigen tests (Section S2).

SARS-CoV-2 testing in Qatar was conducted on a large scale, primarily for non-clinical reasons.^8, 17^ The national mortality database was used to obtain data on all-cause mortality, including deaths occurring at healthcare facilities and elsewhere.^16, 18^ Qatar launched its COVID-19 vaccination program in December 2020 using the BNT162b2 and mRNA-1273 vaccines.^19^ Detailed descriptions of Qatar’s population and national databases have been previously reported.^8, 12, 16–18, 20, 21^

### COVID-19 acute-care and ICU hospitalizations

This study tracked COVID-19 hospitalizations based on a national protocol for COVID-19 case management administered at Hamad Medical Corporation, the national public healthcare provider, and the only authorized entity to provide COVID-19 healthcare in Qatar. A COVID-19 acute-care admission was defined as a record of an acute-care bed admission for an individual who had an active SARS-CoV-2 infection, irrespective of the clinical condition of the admitted individual. Initially, the duration of an active SARS-CoV-2 infection was defined to be 21 days following a SARS-CoV-2-positive test. However, this duration definition was subsequently reduced to 14 days on July 1, 2020 and then to 5 days on October 17, 2022, reflecting changes in policy guidelines in the country.

COVID-19 ICU admission was defined as an ICU bed admission for an individual with an active SARS-CoV-2 infection, and for which the ICU admission clinical team determined that the clinical condition could be related to COVID-19. If an ICU admission for an individual with an active SARS-CoV-2 infection was determined to be unrelated to COVID-19, it was classified as a COVID-19 acute-care admission according to the national protocol.

### Severe, critical, and fatal COVID-19

To ensure stringent and standardized assessment of COVID-19 infection severity, the national protocol for COVID-19 case management required that each hospitalized COVID-19 patient, in an acute-care or ICU bed, undergoes an infection severity assessment every three days using WHO guidelines until discharge or death. Trained medical personnel, independent of study investigators, performed the classification of cases into severe,^6^ critical,^6^ or fatal^7^ COVID-19 categories using individual chart reviews (Section S3). Severe cases were typically hospitalized in acute-care beds, and sometimes in ICU beds out of precaution, while critical cases were always hospitalized in ICU beds.

The incidence of severe COVID-19 cases was defined as the first assessment indicating severe COVID-19 for a given individual during their hospitalization. Similarly, the incidence of critical and fatal COVID-19 cases were defined based on the first assessment indicating critical or fatal COVID-19 during hospitalization. Additionally, if a severe or critical assessment for a newly admitted COVID-19 patient occurred ≥30 days after discharge from the hospital, it was considered a new diagnosis that is independent of the first one.

### Cohort study of incidence of severe, critical, or fatal COVID-19

In addition to examining the link between hospitalization and severity, a national retrospective cohort study was conducted to investigate the incidence of severe, critical, or fatal COVID-19 cases among Qatari citizens between February 28, 2020, and April 21, 2023. The study cohort included all Qataris alive on February 28, 2020, who had at least one record of a SARS-CoV-2 test during the pandemic, which served as a proxy for their presence in Qatar during the study period.

Considering the various testing mandates implemented throughout the pandemic and large-scale routine testing (Section S1),^8, 17^ it is unlikely that any Qatari residing in Qatar did not undergo at least one SARS-CoV-2 test. In total, 2,915,088 SARS-CoV-2 tests were conducted among this cohort of 312,109 Qataris from the onset of the pandemic until April 21, 2023, with an average rate of 9.3 tests per person. With this volume of testing, this cohort is expected to represent virtually the entire Qatari population, a stable affluent population. The size of the cohort is also consistent with the figures reported in the Qatar Census 2020.^22^

We retrieved COVID-19 severity, criticality, and fatality records for every documented SARS-CoV-2 infection or reinfection since the pandemic onset. SARS-CoV-2 reinfection was defined as a documented infection that occurred ≥90 days after an earlier infection, to avoid misclassifying prolonged SARS-CoV-2 positivity as reinfection.^23, 24^ Patients who progressed to severe, critical, or fatal COVID-19 after a documented infection (or reinfection) were classified based on the worst assessment outcome related to that infection (or reinfection), starting with death,^7^ followed by critical disease,^6^ and then severe disease^6^ (Section S3). The date of incidence of the outcome in this analysis was set as the day of the positive SARS-CoV-2 test that documented the infection that progressed into the severe forms of COVID-19.

All individuals were followed from the study start date (February 28, 2020) until any of the following events: documented infection/reinfection associated with fatal COVID-19, or non-COVID-19-related death, or administrative end of follow-up (April 21, 2023). The pandemic was categorized into distinct phases based on the level of SARS-CoV-2 incidence and the predominant variant (Section S4).

### Statistical analysis

The national cohort was characterized by calculating frequency distributions and measures of central tendency. The cumulative incidence of severe, critical, or fatal COVID-19 was defined as the proportion of individuals at risk (that is members of this national cohort) who had at least one episode of severe, critical, or fatal COVID-19 and was estimated using the Kaplan–Meier estimator method.^25^ Likewise, the cumulative incidence of fatal COVID-19 was estimated.

The incidence rate of any severe, critical, or fatal COVID-19 outcome was determined by dividing the number of episodes by the total person-years contributed by all individuals in the cohort. To estimate the incidence rate, we employed a Poisson log-likelihood regression model with the Stata 17.0 *stptime* command, which provided both the incidence rate and its corresponding 95% confidence interval (CI).

Using the same approach, we also estimated the incidence rate of a fatal COVID-19 outcome. These incidence rates were assessed for the entire follow-up period as well as for different phases of the pandemic to investigate temporal trends.

Incidence rate ratios were estimated to compare the incidence of severe, critical, or fatal COVID-19 by sex, 10-year age group, and number of coexisting medical conditions (0, 1, 2, 3, 4, 5, 6+; Section S5). This analysis estimated effects using a multivariable Poisson regression model to adjust simultaneously for the different factors. Interactions were not investigated. Analogously the same analysis was done for only fatal COVID-19. All statistical analyses were conducted using Stata/SE version 17.0 (StataCorp LLC, College Station, TX, USA).

### Oversight

Hamad Medical Corporation and Weill Cornell Medicine-Qatar Institutional Review Boards approved this retrospective study with a waiver of informed consent. The study was reported following the Strengthening the Reporting of Observational Studies in Epidemiology (STROBE) guidelines (Table S1).

## Results

### Acute-care hospitalizations and severe cases

Figure 1A presents the occurrence of COVID-19 acute-care hospitalizations and of severe COVID-19 cases among Qataris on a monthly basis since onset of the pandemic. The data include a total of 6,742 acute-care admissions, with 4,384 occurring during the pre-omicron phase, 843 during the first wave of the omicron variant (BA.1 & BA.2), and 1,515 in the post-first omicron phase.

**Figure 1.**
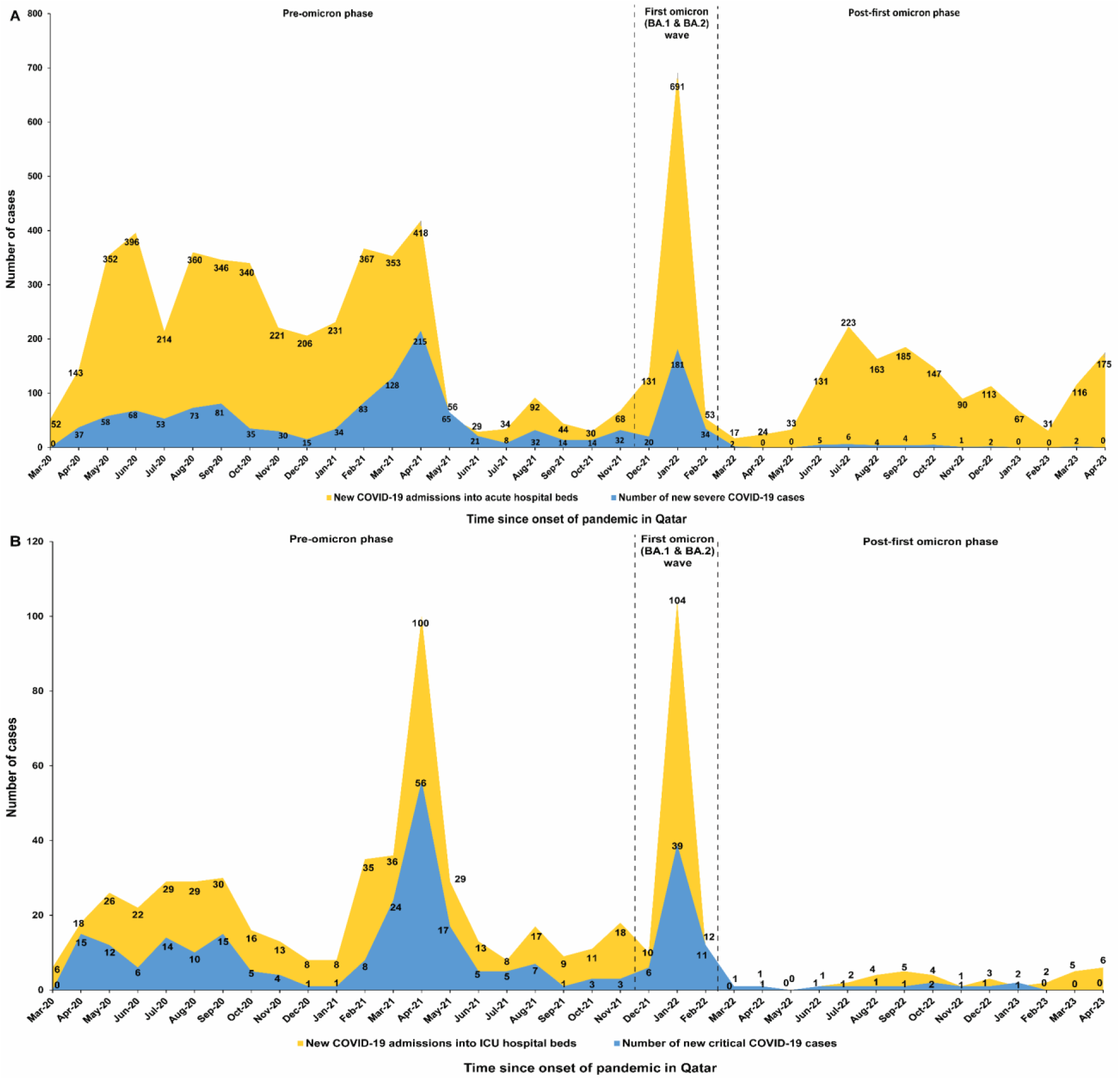
COVID-19 acute-care and ICU bed hospitalizations and COVID-19 severity and criticality among Qataris throughout the pandemic. A) Comparison of the number of new COVID-19 admissions into acute-care hospital beds and the number of new severe COVID-19 cases according to the WHO definition for COVID-19 severity. B) Comparison of the number of new COVID-19 admissions into ICU hospital beds and the number of new critical COVID-19 cases according to the WHO definition for COVID-19 criticality.

Out of the total acute-care admissions, 1,362 cases met the criteria set by the WHO for severe COVID-19 (Figure 1A). Among these severe cases, 1,103 were reported during the pre-omicron phase, 228 during the first omicron wave, and 31 in the post-first omicron phase.

The higher number of acute-care admissions compared to severe cases is attributed to admissions that did not meet the exact definition of severe COVID-19 and hospitalizations with COVID-19 rather than strictly because of COVID-19—incidental hospitalizations or hospitalizations for conditions that COVID-19 may have directly or indirectly worsened. In the very early stage of the pandemic, hospitalization was also utilized for isolation purposes.

### ICU hospitalizations and critical cases

Figure 1B presents the occurrence of COVID-19 ICU hospitalizations and of critical COVID-19 cases among Qataris. The data include a total of 641 ICU admissions, with 485 occurring during the pre-omicron phase, 121 during the first omicron wave, and 35 in the post-first omicron phase.

Out of the total ICU admissions, 281 cases met the criteria set by the WHO for critical COVID-19 (Figure 1B). Among these critical cases, 215 were reported during the pre-omicron phase, 54 during the first omicron wave, and 12 in the post-first omicron phase.

The number of ICU admissions surpassed the count of critical cases, although the difference was not as large as the gap observed between acute-care admissions and severe cases. The gap appears to be attributed to precautionary admissions not meeting the precise criteria for critical COVID-19, and some admissions being with COVID-19 rather than strictly because of COVID-19.

### All-cause and COVID-19 deaths

Figure 2 depicts the occurrence of all-cause deaths and fatal COVID-19 cases among Qataris. The data comprises a total of 2,002 all-cause deaths, with 1,045 occurring during the pre-omicron phase, 192 during the first omicron wave, and 765 in the post-first omicron phase (Figure 2A).

**Figure 2.**
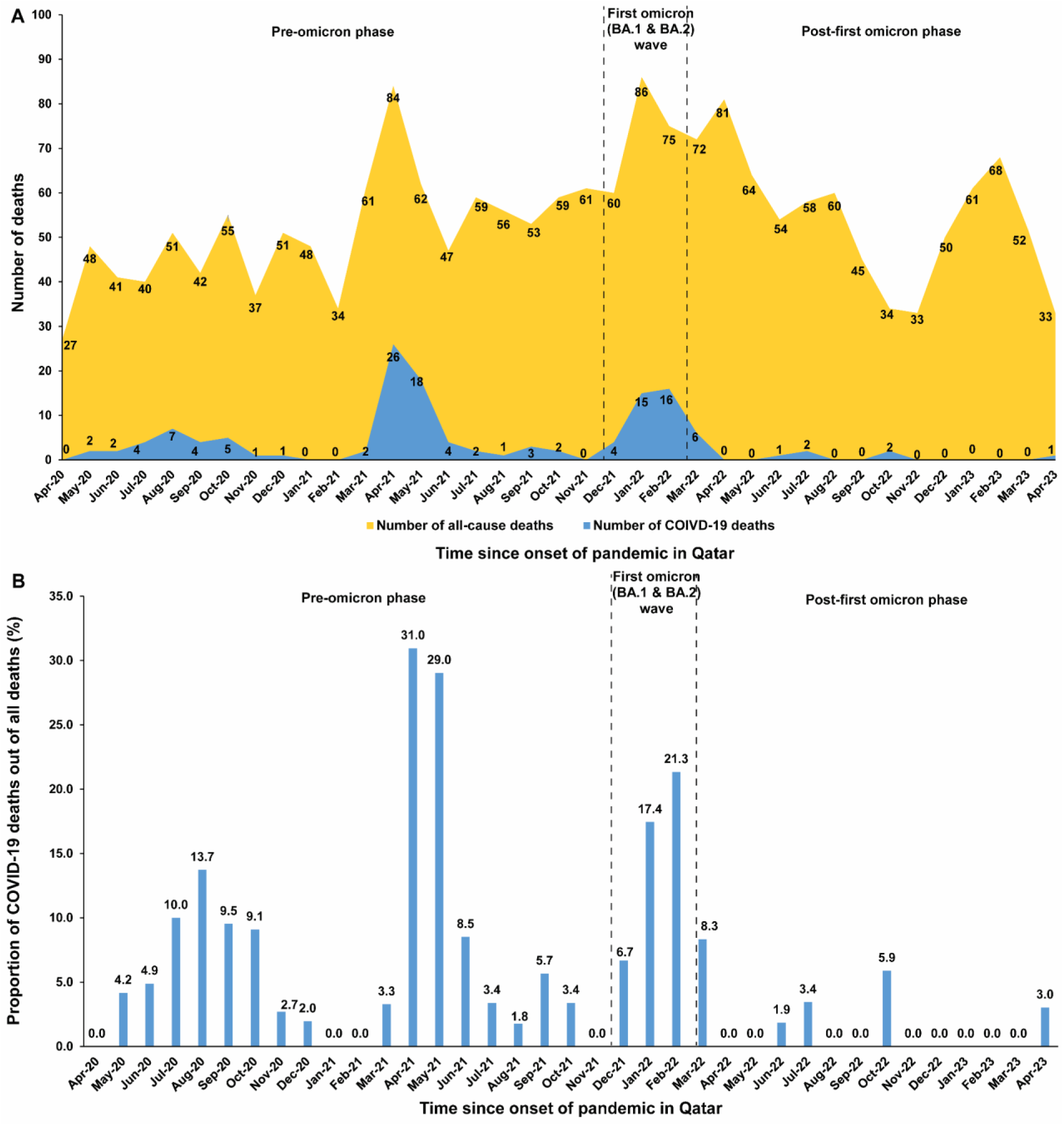
All-cause and COVID-19 deaths among Qataris throughout the pandemic. A) Number of all-cause deaths and of COVID-19 deaths. B) Proportion of COVID-19 deaths out of all-cause deaths.

Among all-cause deaths, 131 cases met the WHO criteria for fatal COVID-19 (Figure 2A). Out of these deaths, 86 were reported during the pre-omicron phase, 33 during the first omicron wave, and 12 in the post-first omicron phase. COVID-19 deaths comprised 6.5% of all-cause deaths. This proportion was highest during the first omicron wave at 17.2% and lowest during the post-first omicron phase at only 1.6% (Figure 2B).

### Incidence rate of severe, critical, or fatal COVID-19

Table 1 provides the baseline characteristics of the participants in the cohort study. Figure 3A visually represents the incidence of SARS-CoV-2 infection, including periods of dominance of various variants throughout the pandemic (dates are found in Section S4).

**Figure 3.**
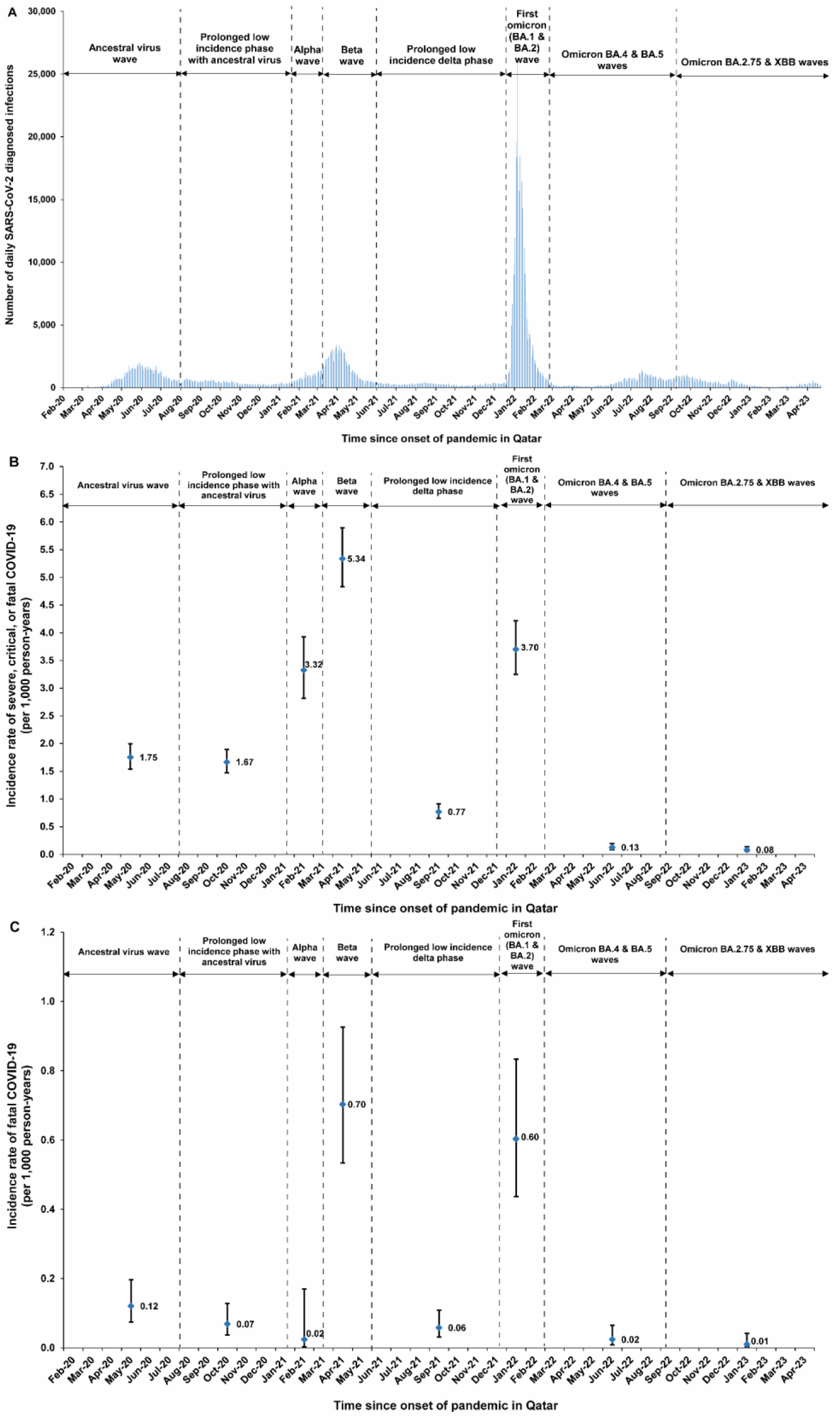
Temporal patterns of infections and severe and fatal COVID-19 during the waves and phases of the pandemic. A) Daily count of newly diagnosed SARS-CoV-2 infections. B) Incidence rate of severe, critical, or fatal COVID-19. C) Incidence rate of fatal COVID-19.

**Table 1.** Baseline characteristics of study participants.

| Characteristics | National Qatari cohort |
| --- | --- |
|  | N=312,109 |
| Median age (interquartile range)—years | 23 (11-39) |
| Age group |  |
| 0-9 years | 74,519 (23.9) |
| 10-19 years | 68,051 (21.8) |
| 20-29 years | 53,644 (17.2) |
| 30-39 years | 43,099 (13.8) |
| 40-49 years | 30,243 (9.7) |
| 50-59 years | 22,401 (7.2) |
| 60-69 years | 13,200 (4.2) |
| 70+ years | 6,952 (2.2) |
| Sex |  |
| Male | 153,606 (49.2) |
| Female | 158,503 (50.8) |
| Number of coexisting medical conditions |  |
| None | 178,865 (57.3) |
| 1 | 71,225 (22.8) |
| 2 | 28,129 (9.0) |
| 3 | 12,547 (4.0) |
| 4 | 8,439 (2.7) |
| 5 | 5,659 (1.8) |
| 6+ | 7,245 (2.3) |

There were 1,396 episodes of severe, critical, or fatal COVID-19 during a total follow-up of 978,041 person-years. The median follow-up time was 3.14 years (interquartile range: 3.14-3.14 years). The majority of infected individuals (99.5%) progressed to severe, critical, or fatal COVID-19 subsequent to the primary infection (as opposed to subsequent to reinfection). The cumulative incidence of severe, critical, or fatal COVID-19 after 3.14 years of follow-up was 0.45% (95% CI: 0.43-0.47%), with two major jumps in incidence during the beta and omicron waves (Figure 4A).

**Figure 4.**
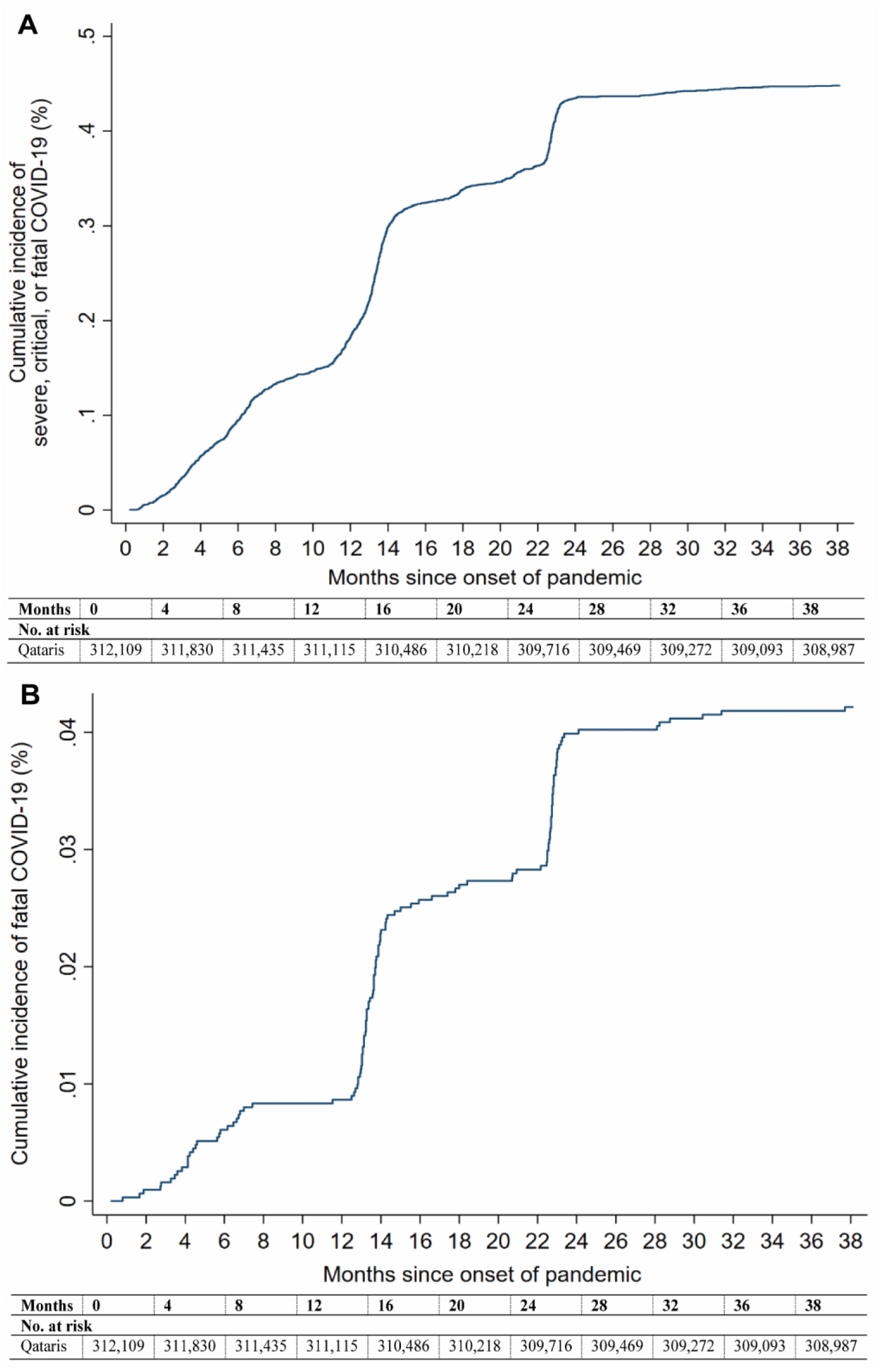
Cumulative incidence of A) severe, critical, or fatal COVID-19 and B) fatal COVID-19 since the onset of the pandemic.

The incidence rate of severe, critical, or fatal COVID-19 was 1.43 (95% CI: 1.35-1.50) per 1,000 person-years throughout the pandemic (Figure 5A). During the pre-omicron phase, the first omicron wave, and these two phases combined, it was 2.01 (95% CI: 1.90-2.13), 3.70 (95% CI: 3.25-4.22), and 2.18 (95% CI: 2.07-2.30) per 1,000 person-years, respectively. In the post-first omicron phase, the incidence rate dropped drastically to 0.10 (95% CI: 0.08-0.14) per 1,000 person-years, a 95.4% reduction compared to earlier phases.

**Figure 5.**
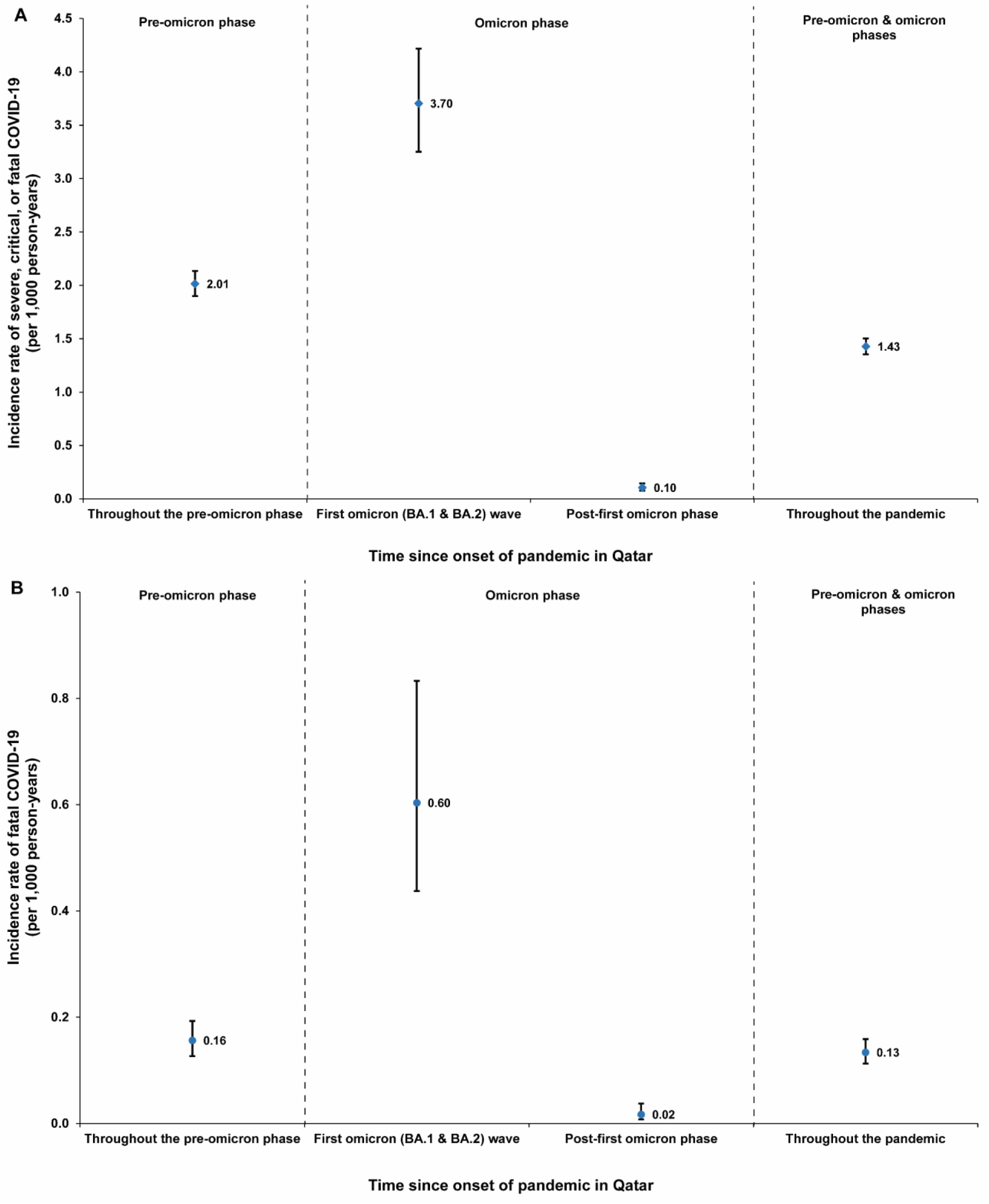
Incidence rates of A) severe, critical, or fatal COVID-19 and B) fatal COVID-19 during the pre-omicron phase, first omicron wave, and post-first omicron phase.

The highest incidence rates were observed during the beta and first omicron waves, while the post-first omicron waves consistently had very low incidence rates (Figure 3B).

### Incidence rate of fatal COVID-19

A total of 131 individuals developed fatal COVID-19 during follow-up, all of which occurred after the primary infection and none during reinfection. The cumulative incidence of fatal COVID-19 after 3.14 years of follow-up was 0.042% (95% CI: 0.036-0.050%), with two major jumps in incidence during the beta and omicron waves (Figure 4B).

The incidence rate of fatal COVID-19 was 0.13 (95% CI: 0.11-0.16) per 1,000 person-years throughout the pandemic (Figure 5B). During the pre-omicron phase, the first omicron wave, and these two phases combined, it was 0.16 (95% CI: 0.13-0.19), 0.60 (95% CI: 0.44-0.83), and 0.20 (95% CI: 0.17-0.24) per 1,000 person-years, respectively. In the post-first omicron phase, the incidence rate dropped drastically to 0.02 (95% CI: 0.01-0.04) per 1,000 person-years, a 90.0% reduction compared to earlier phases.

The highest incidence rates were observed during the beta and first omicron waves, while the post-first omicron waves consistently had very low incidence rates (Figure 3C).

### Associations with severe and fatal COVID-19

The adjusted incidence rate ratio for severe, critical, or fatal COVID-19 was lower in females compared to males, increased exponentially with age, and linearly with the number of coexisting conditions (Figures 6A-C). Similar associations were observed for fatal COVID-19 (Figures 6D-F).

**Figure 6.**
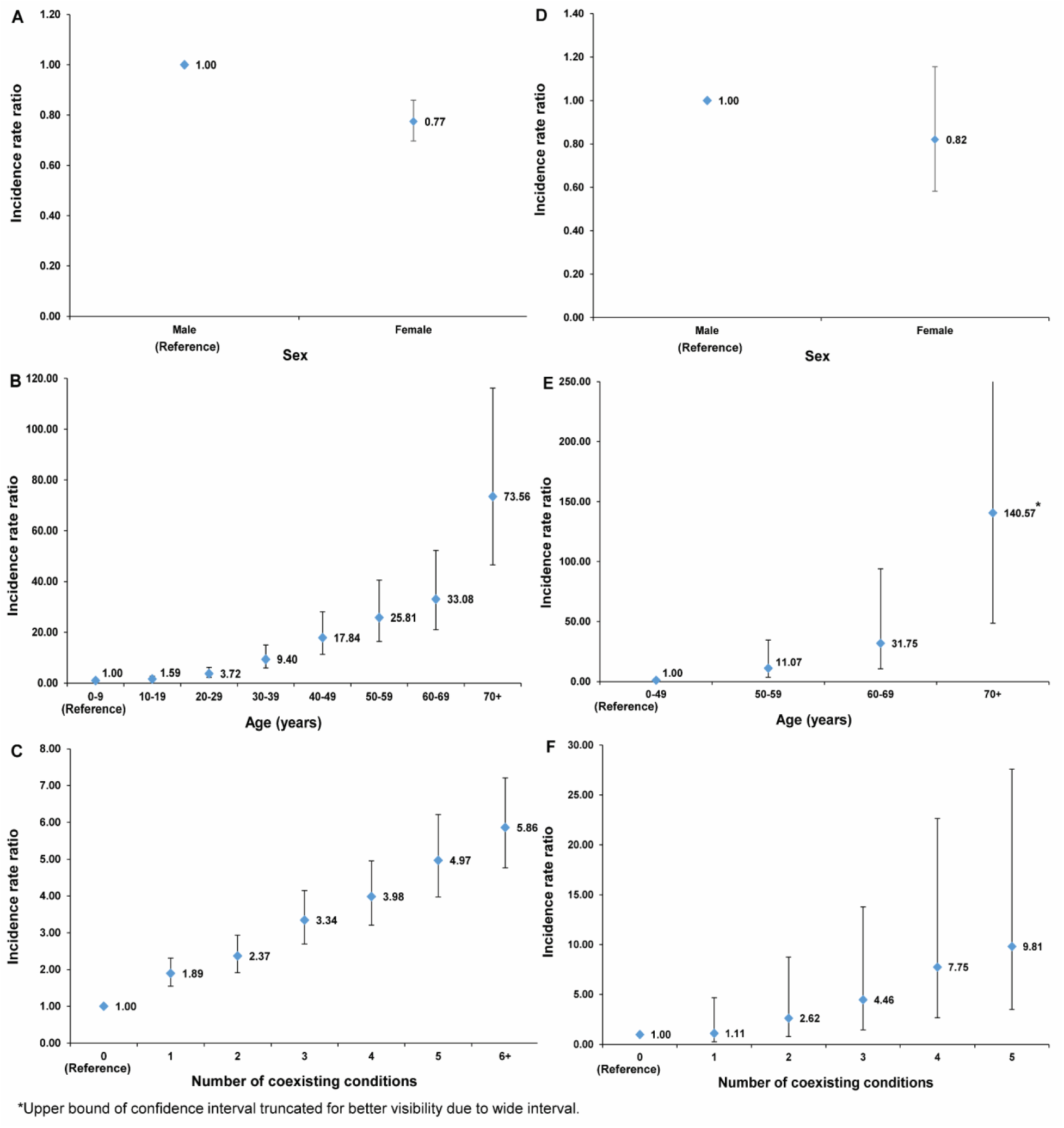
Adjusted incidence rate ratios for severe, critical, or fatal COVID-19 and for only fatal COVID-19 across sex (A and D), age (B and E), and number of coexisting conditions (C and F), respectively.

## Discussion

The end of the first omicron wave marked a turning point in the pandemic, with a 95% drop in severe, critical, or fatal cases of COVID-19 compared to earlier stages. While vaccinations and advancements in case management contributed to reducing severity and fatality gradually over time,^8, 20, 26^ the end of the first omicron wave served as the central turning point.

This turning point is likely attributed to the rapid build-up of natural immunity during the initial omicron wave, which had the highest infection incidence throughout the pandemic. This aligns with earlier studies in Qatar, demonstrating strong protection against severe reinfection among individuals with prior infection and limited waning in protection.^11, 27, 28^ Other factors, such as lower severity in omicron subvariants,^29–32^ the overall decreased infection incidence in the post-first omicron phase, and the forward displacement of deaths of individuals with relatively short life expectancy,^18^ have all also likely contributed to this shift in severity.

A defining aspect of this transition is the near-complete decoupling between infection and severity after the first omicron wave (Figure 3), and between acute-care admissions and severe cases (Figure 1A), as reported elswhere.^33^ Meanwhile there was some decoupling between ICU admissions and critical cases, albeit to a lesser extent (Figure 1B), suggesting ICU admissions as an early warning indicator for changes in COVID-19 severity.

COVID-19 severity varied throughout the pandemic based on the circulating variant and infection incidence, as well as population immunity due to vaccination and previous infections.^11^ The beta wave exhibited the highest severity due to the severity of that variant^34^ and concentrated incidence (Figure 3). The first omicron wave followed with high severity, despite the lower severity of the omicron variant compared to earlier variants.^29, 35^ This was primarily due to a high concentration of infections within a short duration, and perhaps a stretched healthcare system dealing with a large number of severe cases. The introduction of the delta variant, occurring after the large beta wave and the rapid increase in vaccination,^36, 37^ did not result in high severity, due to its low incidence, a consequence of the significant level of pre-omicron vaccine and previous-infection population immunity accumulated in the population at that time.^11^ In the post-first omicron phase, fatality became rare across all omicron subvariant waves, with only 6 recorded cases in the last 12 months (Figure 2A).

This study has limitations. The study examined severe, critical, and fatal cases of COVID-19, according to the WHO criteria. However, the spectrum of COVID-19 severity might have evolved over time, potentially encompassing milder forms that were not captured by the WHO criteria but still required hospitalization.

Timing differences between COVID-19 hospitalization onset and severity assessment were observed. Severity or criticality was determined based on WHO criteria after a longitudinal review of the patient’s case. COVID-19 deaths often occurred weeks after infection, potentially after the wave had subsided. These factors caused a lag between outcome assessment, explaining minor variations in timing. For example, the recorded 6 COVID-19 deaths in the first month of the post-first omicron phase were actually a result of infections during the initial omicron wave (Figure 2A).

The study’s findings are specific to the national population in which it was conducted, making their generalizability to other national populations uncertain. Qatar, with its high human development index,^38^ well-resourced healthcare system,^39^ and young median age (Table 1), has witnessed lower severity and fatality rates of COVID-19.^1, 16, 18^ While these factors contribute to the observed outcomes, the relative decrease in severity and fatality after the first omicron wave is likely to have broader applicability to other national populations.

Documented COVID-19 deaths may not fully capture the total number of fatalities resulting from the virus,^3, 4^ and some deaths may not have been confirmed due to insufficient information to establish COVID-19 as the cause of death. Some infections may have gone undocumented, especially after November 1, 2022, when testing was reduced from 5% to 1% of the population per week.^8, 17, 32^ The study examined severe, critical, and fatal COVID-19 cases that occurred in Qatar. However, there is a possibility of cases occurring outside of Qatar if Qatari individuals traveled abroad for vacation or other reasons. Nevertheless, it is expected that the number of such cases would be minimal, especially considering the restrictions on travel during the pandemic.

In summary, while vaccinations and advancements in case management have contributed to a gradual reduction in severity and fatality over time in Qatar, the end of the first omicron wave marked a significant turning point in the trajectory of severity during this pandemic. The post-first omicron phase exhibited a remarkable 95% reduction in the incidence rate of severe, critical, or fatal cases compared to earlier stages, accompanied by a 90% decrease in the incidence rate of fatal COVID-19. This shift in severity is believed to be driven by the rapid accumulation of natural immunity during the initial omicron wave. Given the limited observed waning in the protection provided by natural immunity against severe reinfection,^11, 28^ it is plausible that the phase of low severity could be sustained as long as the virus does not undergo extensive evolution beyond what has been observed since the introduction of omicron, and the population does not have high rates of comorbidity, which could dangerously exacerbate a repeat infection.

## Data Availability

The dataset of this study is a property of the Qatar Ministry of Public Health that was provided to the researchers through a restricted-access agreement that prevents sharing the dataset with a third party or publicly. The data are available under restricted access for preservation of confidentiality of patient data. Access can be obtained through a direct application for data access to Her Excellency the Minister of Public Health (https:// www.moph.gov.qa/english/OurServices/eservices/Pages/Governmental-HealthCommunication-Center.aspx). The raw data are protected and are not available due to data privacy laws. Aggregate data are available within the paper and its supplementary information.

## Acknowledgments

We acknowledge the many dedicated individuals at Hamad Medical Corporation, the Ministry of Public Health, the Primary Health Care Corporation, Qatar Biobank, Sidra Medicine, and Weill Cornell Medicine-Qatar for their diligent efforts and contributions to make this study possible.

## Author contributions

HC co-designed the study, performed the statistical analyses, and co-wrote the first draft of the article. LJA conceived and co-designed the study, led the statistical analyses, and co-wrote the first draft of the article. PVC conducted viral genome sequencing and designed mass PCR testing to allow routine capture of SGTF variants. PT and MRH conducted the multiplex, real-time reverse-transcription PCR variant screening and viral genome sequencing. HY, AAA-T, and HAK conducted viral genome sequencing. All authors contributed to data collection and acquisition, database development, discussion and interpretation of the results, and to the writing of the manuscript. All authors have read and approved the final manuscript.

## Data availability

The dataset of this study is a property of the Qatar Ministry of Public Health that was provided to the researchers through a restricted-access agreement that prevents sharing the dataset with a third party or publicly. The data are available under restricted access for preservation of confidentiality of patient data. Access can be obtained through a direct application for data access to Her Excellency the Minister of Public Health (https://www.moph.gov.qa/english/OurServices/eservices/Pages/Governmental-HealthCommunication-Center.aspx). The raw data are protected and are not available due to data privacy laws. Aggregate data are available within the paper and its supplementary information.

## Supplementary Appendix

### Section S1. Study population and data sources

Qatar’s national and universal public healthcare system uses the Cerner-system advanced digital health platform to track all electronic health record encounters of each individual in the country, including all citizens and residents registered in the national and universal public healthcare system. Registration in the public healthcare system is mandatory for citizens and residents.

The databases analyzed in this study are data-extract downloads from the Cerner-system that have been implemented on a regular (twice weekly) schedule since onset of the pandemic by the Business Intelligence Unit at Hamad Medical Corporation. Hamad Medical Corporation is the national public healthcare provider in Qatar. At every download all tests, coronavirus disease 2019 (COVID-19) vaccinations, hospitalizations related to COVID-19, and all death records regardless of cause are provided to the authors through .csv files. These databases have been analyzed throughout the pandemic not only for study-related purposes, but also to provide policymakers with summary data and analytics to inform the national response.

Every health encounter in the Cerner-system is linked to a unique individual through the HMC Number that links all records for this individual at the national level. Databases were merged and analyzed using the HMC Number to link all records whether for testing, vaccinations, hospitalizations, and deaths. All deaths in Qatar are tracked by the public healthcare system. All COVID-19-related healthcare was provided only in the public healthcare system. No private entity was permitted to provide COVID-19-related hospitalization. COVID-19 vaccination was also provided only through the public healthcare system. These health records were tracked throughout the COVID-19 pandemic using the Cerner system. This system has been implemented in 2013, before the onset of the pandemic. Therefore, we had all health records related to this study for the full national cohort of Qataris throughout the pandemic. This allowed us to follow each person over time.

Demographic details for every HMC Number (individual) such as sex, age, and nationality are collected upon issuing of the universal health card, based on the Qatar Identity Card, which is a mandatory requirement by the Ministry of Interior to every citizen and resident in the country. Data extraction from the Qatar Identity Card to the digital health platform is performed electronically through scanning techniques.

All severe acute respiratory syndrome coronavirus 2 (SARS-CoV-2) testing in any facility in Qatar is tracked nationally in one database, the national testing database. This database covers all testing in all locations and facilities throughout the country, whether public or private. Every polymerase chain reaction (PCR) test and a proportion of the facility-based rapid antigen tests conducted in Qatar, regardless of location or setting, are classified on the basis of symptoms and the reason for testing (clinical symptoms, contact tracing, surveys or random testing campaigns, individual requests, routine healthcare testing, pre-travel, at port of entry, or other).

Before November 1, 2022, SARS-CoV-2 testing in Qatar was done at a mass scale where about 5% of the population were tested every week.^1, 2^ Based on the distribution of the reason for testing up to November 1, 2022, most of the tests in Qatar were conducted for routine reasons, such as being travel-related, and about 75% of cases were diagnosed not because of appearance of symptoms, but because of routine testing.^1, 2^

Starting from November 1, 2022, SARS-CoV-2 testing was substantially reduced, but still about 1% of the population are tested every week.^3^ All testing results in the national testing database during follow-up in the present study were factored in the analyses of this study.

The first large omicron wave that peaked in January of 2022 was massive and strained the testing capacity in the country.^1, 4–6^ Accordingly, rapid antigen testing was introduced to relieve the pressure on PCR testing. Implementation of this change in testing policy occurred quickly precluding incorporation of reason for testing in a large proportion of the rapid antigen tests for several months. While the reason for testing is available for all PCR tests, it is not available for all rapid antigen tests. Availability of reason for testing for the rapid antigen tests also varied with time.

Rapid antigen test kits are available for purchase in pharmacies in Qatar, but outcome of home-based testing is not reported nor documented in the national databases. Since SARS-CoV-2-test outcomes were linked to specific public health measures, restrictions, and privileges, testing policy and guidelines stress facility-based testing as the core testing mechanism in the population. While facility-based testing is provided free of charge or at low subsidized costs, depending on the reason for testing, home-based rapid antigen testing is de-emphasized and not supported as part of national policy.

Further descriptions of the study population and the national databases were reported previously.^1, 2, 5, 7–10^

### Section S2. Laboratory methods and variant ascertainment

#### Real-time reverse-transcription polymerase chain reaction testing

Nasopharyngeal and/or oropharyngeal swabs were collected for polymerase chain reaction (PCR) testing and placed in Universal Transport Medium (UTM). Aliquots of UTM were: 1) extracted on KingFisher Flex (Thermo Fisher Scientific, USA), MGISP-960 (MGI, China), or ExiPrep 96 Lite (Bioneer, South Korea) followed by testing with real-time reverse-transcription PCR (RT-qPCR) using TaqPath COVID-19 Combo Kits (Thermo Fisher Scientific, USA) on an ABI 7500 FAST (Thermo Fisher Scientific, USA); 2) tested directly on the Cepheid GeneXpert system using the Xpert Xpress SARS-CoV-2 (Cepheid, USA); or 3) loaded directly into a Roche cobas 6800 system and assayed with the cobas SARS-CoV-2 Test (Roche, Switzerland). The first assay targets the viral S, N, and ORF1ab gene regions. The second targets the viral N and E-gene regions, and the third targets the ORF1ab and E-gene regions.

All PCR testing was conducted at the Hamad Medical Corporation Central Laboratory or Sidra Medicine Laboratory, following standardized protocols.

#### Rapid antigen testing

Severe acute respiratory syndrome coronavirus 2 (SARS-CoV-2) antigen tests were performed on nasopharyngeal swabs using one of the following lateral flow antigen tests: Panbio COVID-19 Ag Rapid Test Device (Abbott, USA); SARS-CoV-2 Rapid Antigen Test (Roche, Switzerland); Standard Q COVID-19 Antigen Test (SD Biosensor, Korea); or CareStart COVID-19 Antigen Test (Access Bio, USA). All antigen tests were performed point-of-care according to each manufacturer’s instructions at public or private hospitals and clinics throughout Qatar with prior authorization and training by the Ministry of Public Health (MOPH). Antigen test results were electronically reported to the MOPH in real time using the Antigen Test Management System which is integrated with the national Coronavirus Disease 2019 (COVID-19) database.

#### Classification of infections by variant type

Surveillance for SARS-CoV-2 variants in Qatar is based on viral genome sequencing and multiplex RT-qPCR variant screening^11^ of random positive clinical samples,^2, 12–16^ complemented by deep sequencing of wastewater samples.^14, 17, 18^ Further details on the viral genome sequencing and multiplex RT-qPCR variant screening throughout the SARS-CoV-2 waves in Qatar can be found in previous publications.^1, 2, 4, 8, 12–16, 19–23^

### Section S3. COVID-19 severity, criticality, and fatality classification

Classification of Coronavirus Disease 2019 (COVID-19) case severity (acute-care hospitalizations),^24^ criticality (intensive-care-unit hospitalizations),^24^ and fatality^25^ followed World Health Organization (WHO) guidelines. Assessments were made by trained medical personnel independent of study investigators and using individual chart reviews, as part of a national protocol applied to every hospitalized COVID-19 patient. Each hospitalized COVID-19 patient underwent an infection severity assessment every three days until discharge or death.

#### Severe COVID-19

Severe COVID-19 disease was defined per WHO classification as a SARS-CoV-2 infected person with “oxygen saturation of <90% on room air, and/or respiratory rate of >30 breaths/minute in adults and children >5 years old (or ≥60 breaths/minute in children <2 months old or ≥50 breaths/minute in children 2-11 months old or ≥40 breaths/minute in children 1–5 years old), and/or signs of severe respiratory distress (accessory muscle use and inability to complete full sentences, and, in children, very severe chest wall indrawing, grunting, central cyanosis, or presence of any other general danger signs)”.^24^ Detailed WHO criteria for classifying Severe acute respiratory syndrome coronavirus 2 (SARS-CoV-2) infection severity can be found in the WHO technical report.^24^

#### Critical COVID-19

Critical COVID-19 disease was defined per WHO classification as a SARS-CoV-2 infected person with “acute respiratory distress syndrome, sepsis, septic shock, or other conditions that would normally require the provision of life sustaining therapies such as mechanical ventilation (invasive or non-invasive) or vasopressor therapy”.^24^ Detailed WHO criteria for classifying SARS-CoV-2 infection criticality can be found in the WHO technical report.^24^

#### Fatal COVID-19

COVID-19 death was defined per WHO classification as “a death resulting from a clinically compatible illness, in a probable or confirmed COVID-19 case, unless there is a clear alternative cause of death that cannot be related to COVID-19 disease (e.g. trauma). There should be no period of complete recovery from COVID-19 between illness and death. A death due to COVID-19 may not be attributed to another disease (e.g. cancer) and should be counted independently of preexisting conditions that are suspected of triggering a severe course of COVID-19”. Detailed WHO criteria for classifying COVID-19 death can be found in the WHO technical report.^25^

### Section S4. Phases of the COVID-19 pandemic

The pandemic was categorized into distinct phases based on the level of SARS-CoV-2 incidence and the predominant variant. These phases included the ancestral virus wave (February 28, 2020 - July 31, 2020),^7^ a prolonged low incidence phase with the ancestral virus (August 1, 2020 - January 17, 2021),^2, 26^ the alpha wave (January 18, 2021 - March 7, 2021),^27^ the beta wave (March 8, 2021 - May 31, 2021),^28^ a prolonged low incidence delta phase (June 1, 2021 - December 18, 2021),^19, 29^ the first (BA.1 & BA.2) omicron wave (December 19, 2021 - February 28, 2022),^6^ the omicron BA.4 & BA.5 wave (March 1, 2022 - September 9, 2022),^22^ and the omicron BA.2.75 & XBB waves (September 10, 2022 - April 21, 2023).^3^

### Section S5. Comorbidity classification

Comorbidities were ascertained and classified based on the ICD-10 codes as recorded in the electronic health record encounters of each individual in the Cerner-system national database that includes all citizens and residents registered in the national and universal public healthcare system. The public healthcare system provides healthcare to the entire resident population of Qatar free of charge or at heavily subsidized costs, including prescription drugs.

All encounters for each individual were analyzed to determine the comorbidity classification for that individual, as part of a recent national analysis to assess healthcare needs and resource allocation. The Cerner-system national database includes encounters starting from 2013, after this system was launched in Qatar. As long as each individual had at least one encounter with a specific comorbidity diagnosis since 2013, this person was classified with this comorbidity.

Individuals who have comorbidities but never sought care in the public healthcare system, or seek care exclusively in private healthcare facilities, were classified as individuals with no comorbidity due to absence of recorded encounters for them.

**Table S1.**
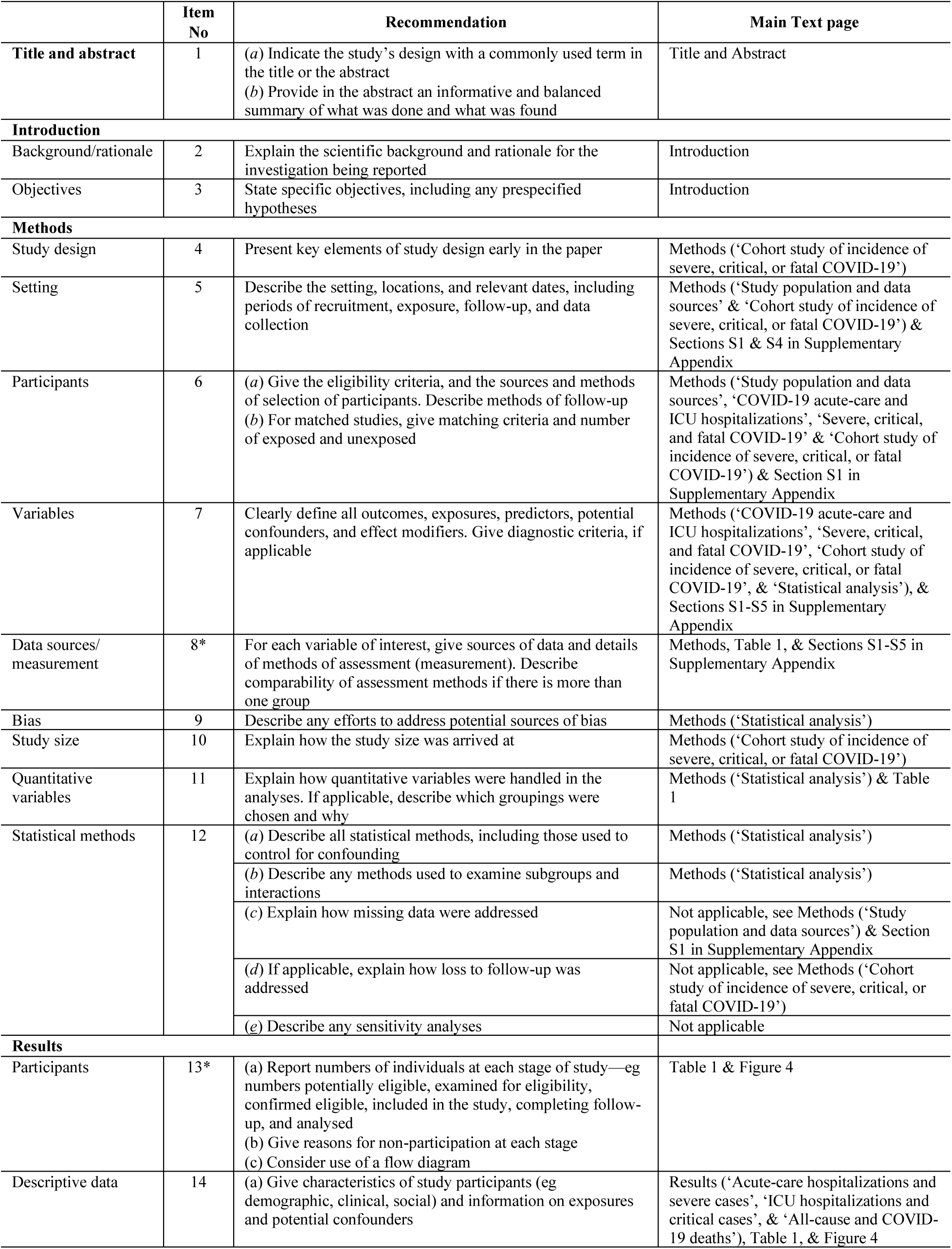
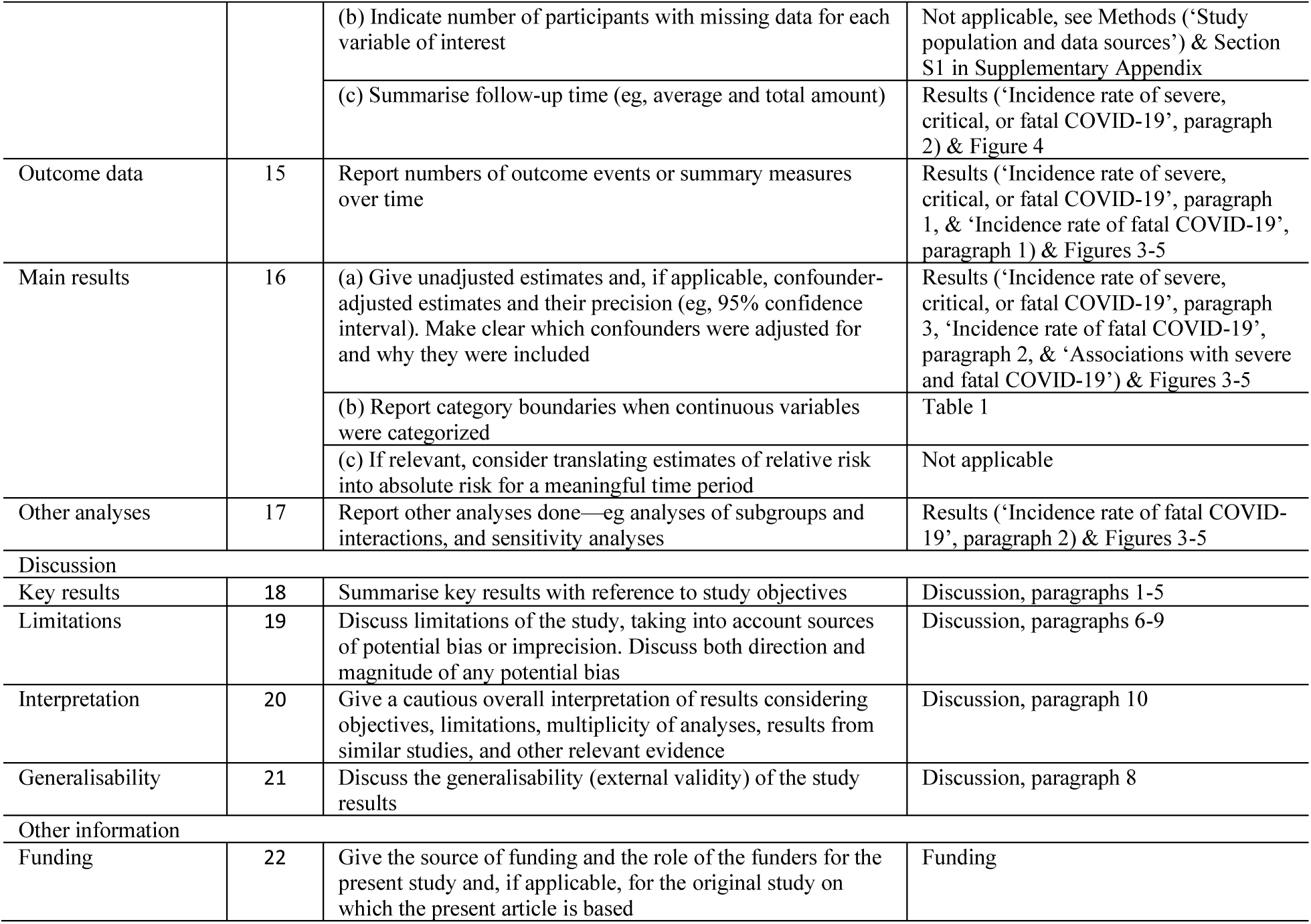
STROBE checklist for cohort studies.

|  | Item No | Recommendation | Main Text page |
| --- | --- | --- | --- |
| Title and abstract | 1 | (a) Indicate the study’s design with a commonly used term in the title or the abstract<br>(b) Provide in the abstract an informative and balanced summary of what was done and what was found | Title and Abstract |
| Introduction |  |  |  |
| Background/rationale | 2 | Explain the scientific background and rationale for the investigation being reported | Introduction |
| Objectives | 3 | State specific objectives, including any prespecified hypotheses | Introduction |
| Methods |  |  |  |
| Study design | 4 | Present key elements of study design early in the paper | Methods (‘Cohort study of incidence of severe, critical, or fatal COVID-19’) |
| Setting | 5 | Describe the setting, locations, and relevant dates, including periods of recruitment, exposure, follow-up, and data collection | Methods (‘Study population and data sources’ & ‘Cohort study of incidence of severe, critical, or fatal COVID-19’) & Sections S1 & S4 in Supplementary Appendix |
| Participants | 6 | (a) Give the eligibility criteria, and the sources and methods of selection of participants. Describe methods of follow-up<br>(b) For matched studies, give matching criteria and number of exposed and unexposed | Methods (‘Study population and data sources’, ‘COVID-19 acute-care and ICU hospitalizations’, ‘Severe, critical, and fatal COVID-19’ & ‘Cohort study of incidence of severe, critical, or fatal COVID-19’) & Section S1 in Supplementary Appendix |
| Variables | 7 | Clearly define all outcomes, exposures, predictors, potential confounders, and effect modifiers. Give diagnostic criteria, if applicable | Methods (‘COVID-19 acute-care and ICU hospitalizations’, ‘Severe, critical, and fatal COVID-19’, ‘Cohort study of incidence of severe, critical, or fatal COVID-19’, & ‘Statistical analysis’), & Sections S1-S5 in Supplementary Appendix |
| Data sources/measurement | 8* | For each variable of interest, give sources of data and details of methods of assessment (measurement). Describe comparability of assessment methods if there is more than one group | Methods, Table 1, & Sections S1-S5 in Supplementary Appendix |
| Bias | 9 | Describe any efforts to address potential sources of bias | Methods (‘Statistical analysis’) |
| Study size | 10 | Explain how the study size was arrived at | Methods (‘Cohort study of incidence of severe, critical, or fatal COVID-19’) |
| Quantitative variables | 11 | Explain how quantitative variables were handled in the analyses. If applicable, describe which groupings were chosen and why | Methods (‘Statistical analysis’) & Table 1 |
| Statistical methods | 12 | (a) Describe all statistical methods, including those used to control for confounding | Methods (‘Statistical analysis’) |
|  |  | (b) Describe any methods used to examine subgroups and interactions | Methods (‘Statistical analysis’) |
|  |  | (c) Explain how missing data were addressed | Not applicable, see Methods (‘Study population and data sources’) & Section S1 in Supplementary Appendix |
|  |  | (d) If applicable, explain how loss to follow-up was addressed | Not applicable, see Methods (‘Cohort study of incidence of severe, critical, or fatal COVID-19’) |
|  |  | (e) Describe any sensitivity analyses | Not applicable |
| Results |  |  |  |
| Participants | 13* | (a) Report numbers of individuals at each stage of study—eg numbers potentially eligible, examined for eligibility, confirmed eligible, included in the study, completing follow-up, and analysed<br>(b) Give reasons for non-participation at each stage<br>(c) Consider use of a flow diagram | Table 1 & Figure 4 |
| Descriptive data | 14 | (a) Give characteristics of study participants (eg demographic, clinical, social) and information on exposures and potential confounders | Results (‘Acute-care hospitalizations and severe cases’, ‘ICU hospitalizations and critical cases’, & ‘All-cause and COVID-19 deaths’), Table 1, & Figure 4 |
|  |  | (b) Indicate number of participants with missing data for each variable of interest | Not applicable, see Methods ('Study population and data sources') & Section S1 in Supplementary Appendix |
|  |  | (c) Summarise follow-up time (eg, average and total amount) | Results ('Incidence rate of severe, critical, or fatal COVID-19', paragraph 2) & Figure 4 |
| Outcome data | 15 | Report numbers of outcome events or summary measures over time | Results ('Incidence rate of severe, critical, or fatal COVID-19', paragraph 1, & 'Incidence rate of fatal COVID-19', paragraph 1) & Figures 3-5 |
| Main results | 16 | (a) Give unadjusted estimates and, if applicable, confounder-adjusted estimates and their precision (eg, 95% confidence interval). Make clear which confounders were adjusted for and why they were included | Results ('Incidence rate of severe, critical, or fatal COVID-19', paragraph 3, 'Incidence rate of fatal COVID-19', paragraph 2, & 'Associations with severe and fatal COVID-19') & Figures 3-5 |
|  |  | (b) Report category boundaries when continuous variables were categorized | Table 1 |
|  |  | (c) If relevant, consider translating estimates of relative risk into absolute risk for a meaningful time period | Not applicable |
| Other analyses | 17 | Report other analyses done—eg analyses of subgroups and interactions, and sensitivity analyses | Results ('Incidence rate of fatal COVID-19', paragraph 2) & Figures 3-5 |
| Discussion |  |  |  |
| Key results | 18 | Summarise key results with reference to study objectives | Discussion, paragraphs 1-5 |
| Limitations | 19 | Discuss limitations of the study, taking into account sources of potential bias or imprecision. Discuss both direction and magnitude of any potential bias | Discussion, paragraphs 6-9 |
| Interpretation | 20 | Give a cautious overall interpretation of results considering objectives, limitations, multiplicity of analyses, results from similar studies, and other relevant evidence | Discussion, paragraph 10 |
| Generalisability | 21 | Discuss the generalisability (external validity) of the study results | Discussion, paragraph 8 |
| Other information |  |  |  |
| Funding | 22 | Give the source of funding and the role of the funders for the present study and, if applicable, for the original study on which the present article is based | Funding |

